# Trajectories of imitation skills in preschoolers with Autism Spectrum Disorders

**DOI:** 10.1101/2021.03.24.21254258

**Authors:** Irène Pittet, Nada Kojovic, Martina Franchini, Marie Schaer

**Author notes:** (I.P.); (N.K.); (M.S.). (M.F.).

## Abstract

Imitation skills play a crucial role in social cognitive development from early childhood. Many studies have shown a deficit in imitation skills in children with Autism Spectrum Disorders (ASD). Little is known about the development of imitation behaviors in children with ASD. This study aims to measure the trajectories of early imitation skills in preschoolers with ASD and how these skills impact other areas of early development. For this purpose, we assessed imitation, language and cognition skills in 177 children with ASD and 43 typically developing children (TD) aged 2 to 5 years old, 126 of which were followed longitudinally, yielding a total of 396 time-points. Our results confirmed the presence of an early imitation deficit in toddlers with ASD compared to TD children. The study of the trajectories showed that these difficulties were marked at the age of two years, and gradually decreased until the age of 5 years old. Imitation skills were strongly linked with cognitive, language skills and level of symptoms in our ASD group at baseline. Moreover, the imitation skills at baseline were predictive of the language gains a year later in our ASD group. Using a data-driven clustering method, we delineated different developmental trajectories of imitation skills within the ASD group. The clinical implications of the findings are discussed, particularly the impact of an early imitation deficit on other areas of competence of the young child.

## Background

Imitation skills play a crucial role in early development and are considered an essential component of social and cognitive development during the first years of life. Indeed, newborns begin to imitate tongue profusion and mouth opening after only a few hours of life (Heimann & Tjus, 2019; Meltzoff & Moore, 1989). Later, infants begin to imitate vocalisations, actions on objects and gestures. During the two first years of life, children gradually increase their imitative behaviors both in frequency and complexity, along with the development of their cognitive and social abilities (Vivanti & Hamilton, 2014). Children imitate their siblings, peers and parents at every moment to learn new abilities (Howe et al., 2018; Zmyj & Seehagen, 2013). Imitation is an essential lever for learning of new motor skills (Pfeifer et al., 2008), and cognitive skills (Hurley & Chater, 2005; Strid et al., 2006; Vivanti et al., 2013). Besides the role in learning, imitation has an important social function (Uzgiris, 1981, as cited in Ingersoll, 2008). From the earliest moments, caregivers and infants engage in reciprocal imitation during face-to-face interactions. Through this choreography of imitating each other’s vocalisations and facial expressions they will share social interest, affective states, take turns and build a fundament for the development of complex social skills. Indeed, the literature has amply demonstrated that imitation plays a vital role in the development of social communication (Hanika & Boyer, 2019; Toth et al., 2006).

The disruption in early imitation skills can have a tremendous repercussion on various developmental aspects, especially socio-communicative development that is altered in autism. Autism Spectrum Disorder (ASD) is a neurodevelopmental disorder characterized by impairments in social communication, interaction and restricted patterns of interests and activities (American Psychiatric Association, 2013). Indeed, a large body of literature has highlighted broad imitation deficits in children with ASD (Girardot et al., 2009; McDuffie et al., 2007; Plavnick & Hume, 2014; Rogers et al., 1996; Thurm et al., 2007; Vivanti & Hamilton, 2014; Zwaigenbaum et al., 2005) using various imitation measures (Vivanti & Hamilton, 2014). Early imitation deficits were linked to difficulties in social communication skills, such as language (Ingersoll & Meyer, 2011; Poon et al., 2012), joint attention (Villalobos et al., 2005) and social interaction difficulties (Ingersoll, 2008). In addition, such a deficit could hinder the effectiveness of intervention methods for children with ASD, as many of them rely upon imitation to teach new abilities (Dawson et al., 2010; Delano, 2007; Espanola Aguirre & Gutierrez, 2019). Given the critical role that the imitation skills play in typical development, precise characterization of the deficit in this domain in ASD is of utmost importance.

Even though imitation difficulties in ASD are relatively well documented, the developmental trajectories of imitation have received less attention to date in this population. Prospective studies following infants at risk of developing ASD report difficulties in imitation skills in children later diagnosed with ASD from 12 months of age. For example, Young and colleagues (2011) conducted a longitudinal study with 248 children between 12 and 24 months of age and found a delayed imitation development in children who developed ASD by age 3 years compared to the TD group. Likewise, Poon and colleagues (2012) found that infants of 9–12 months of age who are later diagnosed with ASD show a considerable delay in imitation skills. These results have led some research groups to explore if imitation could be used for screening ASD in high-risk siblings. Rowberry and colleagues (2015), for instance, found that poor imitation reported by parents in the First Year Inventory (FYI) (Baranek, Watson, Crais, & Reznick, 2003) can be an early marker of ASD in 12-months-old high-risk siblings. The results of the various studies cited above highlight this imitative deficit in young children with ASD and the importance of increasing and detailing knowledge about the development of this crucial skill and the consequences of such difficulties.

The present study aims to compare the developmental trajectories of imitative skills between preschoolers with ASD and a group of matched TD children, using a total of 396 longitudinal assessments collected between the age of 2 and 5 years old. Based on previous literature (Girardot et al., 2009; McDuffie et al., 2007; Plavnick & Hume, 2014; Rogers et al., 1996; Thurm et al., 2007; Vivanti & Hamilton, 2014; Zwaigenbaum et al., 2005), we expected a delayed development of imitation skills in children with ASD in comparison to TD children. Second, we investigated how imitation skills related to the level of symptoms, cognitive and communication skills in the ASD group at baseline and after one year to explore the consequences of this deficit on the different development areas. We hypothesized that a deficit in imitation skills is linked with severity of symptoms and difficulties in developmental domains at the baseline and one year after (Ingersoll & Meyer, 2011; Poon et al., 2012). Finally, we explored the extent to which there could be different imitation trajectories inside the ASD group, as this population is characterized by a great heterogeneity of profiles and manifestations (Wiggins et al., 2012).

## Material & methods

### Participants

The participants in this study were recruited as part of the Geneva Autism Cohort (Franchini & al., 2016, 2017, 2018; Kojovic & al., 2019; Robain & al., 2020; Sperdin & al., 2018). The University’s Institutional Review Board approved this study. All families provided written informed consent to participate. For the current study, a total of 220 participants were included, 177 with ASD and 43 typically developing children (TD) (see *Table 1* and *Table S1* in *Supplementary Material*s). Among these, 126 were followed longitudinally, yielding a total of 396 time-points (78 subjects had 2, 46 had 3, and 2 participants had 4 time-points), each time-point was separated by 12 months, except for two children for whom the two time-points are separated by 24 months (see *Figure 1*). The inclusion criteria for ASD group was a clinical diagnosis of autism according to the DSM-5 criteria (American Psychiatric Association, 2013). The clinical diagnosis was confirmed by the scores exceeding the cut-off for autism at the ADOS-G (Lord et al., 2000) or ADOS-2 (Lord & al., 2012) (see *Measures* for more details). Children from the TD group had no developmental concerns, no first or second-degree family member diagnosed with autism, and were all administered the ADOS diagnostic test to exclude the presence of ASD symptoms (see *Table 1*). Participants were aged between 2.0 and 5.0 years, and the two groups did not differ by age (*U* = 351, *p* = 0.429) nor sex (*p* = 0.156).

**Figure 1.**
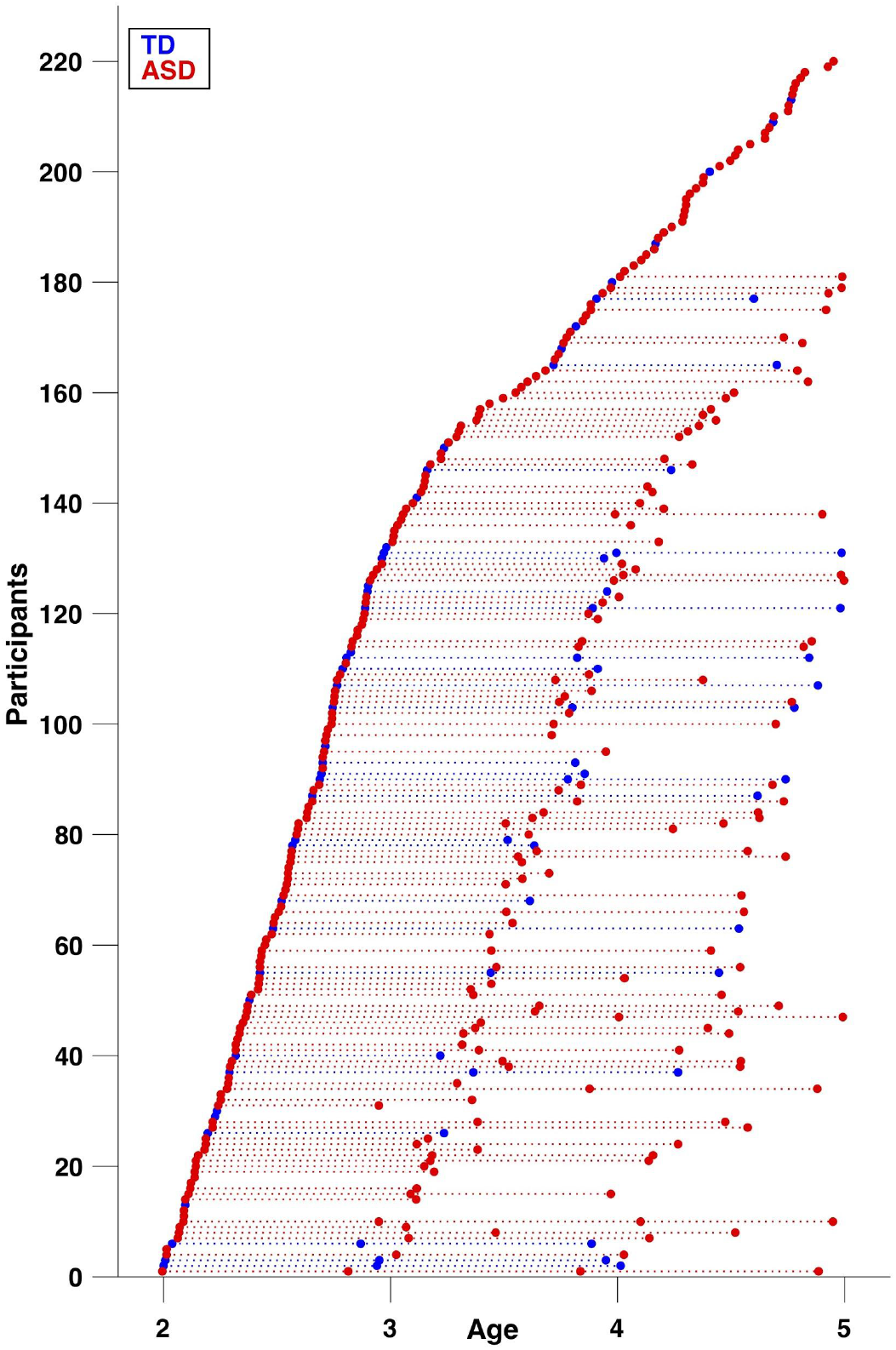
Data distribution available for each group from our longitudinal study. Circles (red = ASD, blue = TD) denote visits/time-points, repeated visits of the same subject are connected with a dotted line.

**Table 1.** Statistical Comparison Between Children With ASD and Children with TD at Baseline in Terms of Demographic, Clinical, Cognitive Features

|  | ASD - Mean (SD) *n* = 177 | TD - Mean (SD) *n* = 43 | *p*-value* |
| --- | --- | --- | --- |
| Sex | 24 ♀ / 153 ♂ | 10 ♀ / 33 ♂ | 0.156<sup>1</sup> |
| Age (years old) | 3.09 (0.8) | 2.93 (0.7) | 0.429<sup>2</sup> |
| ADOS Total symptom severity | **7.7 (1.8)** | **1.02 (1.2)** | **<0.001<sup>2</sup>** |
| PEP-3 |  |  |  |
| VPC (raw score) | **28.47 (15)** | **45 (9.4)** | **<0.001<sup>2</sup>** |
| EL (raw score) | **9.22 (10.2)** | **25.77 (8.8)** | **<0.001<sup>2</sup>** |
| RL (raw score) | **13.41 (10.8)** | **30.57 (5.2)** | **<0.001<sup>2</sup>** |
| VABS-II Adaptive Behavior Composite | **77.61 (11.5)** | **102.9 (8.5)** | **<0.001<sup>2</sup>** |

*Note.* ADOS = Autism Diagnosis Observation Schedule ; PEP-3 = PsychoEducational Profile, 3rd edition ; VPC = Verbal and Preverbal Cognition ; EL = Expressive language ; RL = Receptive language ; VABS-II = Vineland Adaptive Behavior Scales, 2nd edition.
\* *p* value of *Fisher's exact test*<sup>1</sup> and *Mann-Whitney tests*<sup>2</sup> of differences between the ASD and TD groups. Significant results are shown in bold.

## Measures

### Imitation

For the measure of imitation skills we used the Visuo-Motor Imitation scale of the PEP-3 (Schopler & al., 2005) (for a detailed description of this test please refer to Supplementary Materials). This scale includes 10 items of imitation, assessed on a 3 point scale (0 = absence of imitation, 1 = equivoque, partial or prompted imitation, 2 = complete imitation of the skill). Raw scores range between 0 and 20, which allows enough variability to highlight the heterogeneity of imitation profiles.

The scale comprises the imitation of familiar actions and imitation with explicit instruction. Vivanti & Hamilton (2014) distinguished two types of imitation that we find in this scale : *true imitation* (copying both the mean and the goal of the action) and *stimulus enhancement* (perform a specific action on a stimulus after having been made attentive to this stimulus by another person). For instance, one item of the PEP-3 is to organize a fake birthday party to explore imitation of daily actions like eating etc. (*stimulus enhancement*). For another one, the examiner touches her/his nose and says “Now it’s your turn” (*true imitation* of meaningless actions).

### Level of symptoms of autism

Each participant was assessed with the Autism Diagnosis Observation Schedule, 2nd edition (Lord et al., 2012) or Autism Diagnosis Observation Schedule, Generic (ADOS-G; Lord et al., 2000). For subjects who underwent the earlier ADOS version (ADOS-G) assessment, the scores were transformed according to the revised ADOS algorithm (Gotham et al., 2009) to ensure comparability with ADOS-2. The ADOS Calibrated Severity Score (ADOS-CCS; Gotham et al., 2009), a measure of symptom severity relatively independent of language levels and age, was then used for all subjects included in this study.

## Developmental functioning

### Mullen Scale of Early Learning

Developmental functioning of all children included in the present study was assessed with the Psychoeducational Profile, 3rd edition (Schopler et al., 2005). Nevertheless, as our aim was to test the relation between the imitation skills (VMI subscale of PEP-3) and the developmental functioning, we decided to use another measure than the PEP-3 to estimate the level of developmental functioning. We thus used the Mullen Scale of Early Learning-MSEL (Mullen, 1995), which was available for 133 out of the 177 children with ASD included in this study (this measure was indeed added at a later point in our cohort protocol and was thus missing for the data collected earlier).

The MSEL scale is developed for the assessment of typically developing children consists of 5 domains with items organized according to developmental stages and their level of difficulty : Gross Motor (GM), Visual Reception (VR), Fine Motor (FM), Receptive Language (RL) and Expressive Language (EL). This assessment yields a global composite score (Early Learning Composite ; ELC) by adding the four cognitive subscales (VR, FM, RL, EL), and has a mean of 100 and standard deviation of 15. For the global developmental functioning in this study, we considered the standard composite score (ELC). For the measure of individual domains (Receptive and Expressive language), we used the measure of age equivalents (AE). We opted for the AE scores as the standard T scores of the MSEL subscales show poor sensitivity in the lower functioning end of the continuum (floor of 20), and an important number of our participants had this lowest score.

## Statistical Analyses

### Between-group analyses (ASD vs TD)

#### Group comparison of imitation skills

To compare imitation skills at baseline between children with ASD (*n* = 177) and TD children (*n* = 43), we conducted a *t*-test using the raw scores obtained from the Visuo-Motor Imitation scale of PEP-3 (Schopler & al., 2005).

#### Imitation trajectories over time

To compare trajectories of imitation skills over time between ASD and TD children, we used mixed model regression analysis applied on raw imitation scores including all available time points (total of 396 time-points, *N*=220, *n*^*ASD*^=177, *n*^*TD*^=43). This model is particularly adapted for dealing with nested data, such as multiple time points, and has been applied in our previous studies involving a variable number of time points (Franchini & al., 2018; Maeder & al., 2016; Mancini & al., 2019; Mutlu & al., 2013; Schneider & al., 2014). This analysis estimates developmental trajectories by fitting random-slope models to the data, taking into account both within-subject and between-subject effects. Different models (constant, linear, quadratic, or cubic) were fitted using the *nlmefit* function in MATLAB R2011b (MathWorks) for each variable. A Bayesian Information Criterion (BIC)-based model selection method was then employed.

#### Within-group analyses (ASD group)

##### Relation of imitation to the symptom severity and developmental levels at baseline

To explore how an imitation deficit is related to difficulties in other areas of development, we used the cross-sectional data obtained from the ASD group for whom the Mullen Scale of Early Learning was available (*n* = 133, 113 males / 20 females, age range: 2.0 -4.8 years old). The association of imitation with the level of autistic symptoms (CSS ADOS), global cognitive functioning (the Early Learning Composite of MSEL) and more specifically, communication skills (age equivalents on Receptive and Expressive Language subscales of MSEL) was assessed using spearman’s correlations. Results were considered significant at the level of *p* < .0125 (after applying the Bonferroni correction for multiple comparisons).

##### Imitation at baseline as a predictor of symptom severity and developmental levels

We then explored the predictive power of the early imitative skills at baseline on the evolution of autistic symptoms and developmental domains in our ASD group one year after the initial measurement. More specifically, we tested how the imitation predicted the symptom change, change in cognitive skills and communication. To obtain a measure of change, we subtracted the scores at time-point 2 from the ones at time-point 1 (the ADOS-CSS for the level of symptoms, the standard score for the cognitive skills and the age equivalents for communication skills). We then used Spearman partial correlations to assess the relation between imitation skills at baseline and subsequent change in symptoms of autism, cognitive and communication levels a year later, while controlling for the baseline levels.

##### Imitation trajectories within the ASD group

Given the tremendous heterogeneity of the ASD phenotype (Wiggins et al., 2012), we wanted to better understand the diversity of imitation profiles and their evolution in the sample of children with ASD for whom we had longitudinal data (102 subjects). To this end, we used a K-means clustering method, which allows grouping together subjects similar throughout multiple dimensions (i.e. imitation skills at different ages) (Bair, 2013; Sandini et al., 2018). We employed a two-step approach: First, to distinguish between children starting at different levels of imitation skills we employed a K-means clustering on raw scores at the first time-point. To obtain an optimal cluster number among several solutions (2-6 clusters), we used a silhouette approach (Rousseeuw, 1987). Second, we applied a K-means clustering on the slopes of imitation skills by age in the subgroup of children with initially lower levels of imitation skills to test for divergent trajectories over time.

## Results

### Between-group analyses (ASD vs TD)

#### Group comparison of imitation skills

Using a cross-sectional sample (*N* = 220, 177 children with ASD, 43 with TD, age range: 2.0 -5.0 years old), we were able to show that the two groups differed significantly in their imitations skills (*U* = 1270, p < 0.001). Indeed, children with ASD had markedly lower imitation skills than typically developing children (see *Figure 2*).

**Figure 2.**
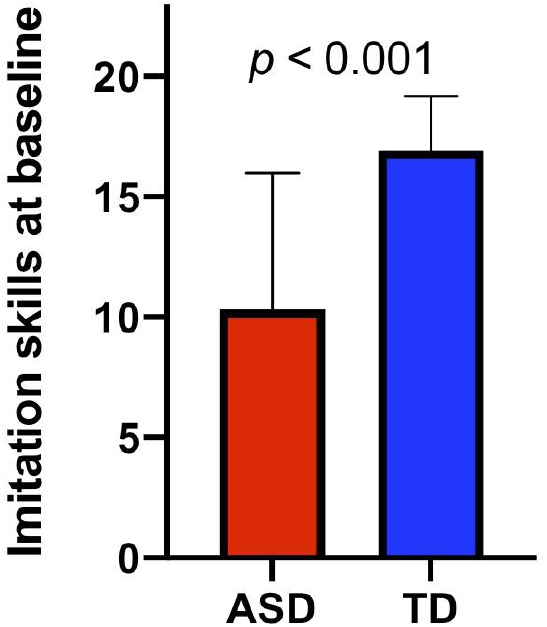
Group comparison of imitation skills obtained from Visuo-Motor Imitation scale of PEP-3 (raw scores).

#### Imitation trajectories over time

Using all available visits for our cross-sectional sample (total of 396 time-points, *N*=220, *n*^*ASD*^=177, *n*^*TD*^=43) we tested the between-group differences in trajectories of imitation. As shown in *Figure 3*, the group trajectories in imitation skills estimated using mixed-models were significantly different (group effect : *p* < .001 ; interaction : *p* < .001). The best fitted model was of the 2nd order, meaning that the relation of the imitation with age was quadratic. Indeed, the TD children showed good imitation skills at two years old with only a slight improvement thereafter. In contrast, at the group level, children with ASD present markedly low imitation skills at a young age with substantial improvement between the ages of two and five (see *Figure 3*). However, we also observe that trajectories of children with ASD are very heterogeneous and this point is addressed in our later analyses.

**Figure 3.**
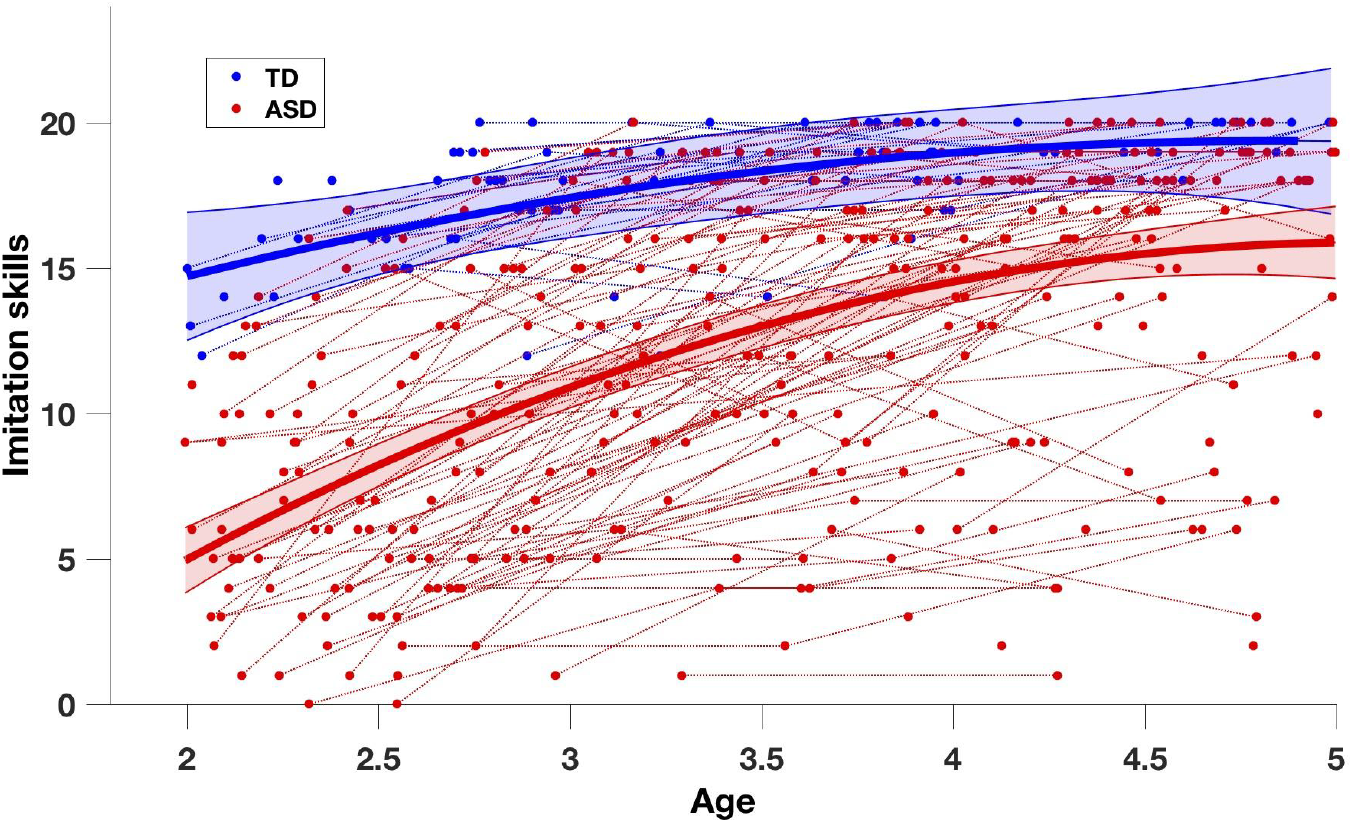
Imitation trajectories over time in the ASD and TD groups. The trajectory on the group level (solid lines, red=ASD, blue=TD) is obtained using mixed-effect models (Mancini & al., 2019; Mutlu & al., 2013). The 95% confidence interval of estimated group-level trajectory is represented in colored bands. Repeated time-points of the same subjects are connected with dotted lines.

## Results within the ASD group

### Relation of imitation with level of symptoms and developmental domains at baseline

Within our ASD group for whom the Mullen Scales of Early Learning was available (*n* = 133, 113 males / 20 females, age range: 2.0 -4.8 years old), imitation skills were moderately negatively related to the level of symptoms (*r*^*s*^ = -0.583, *p* < .001), as the children who show the most of symptoms are also those with more imitation difficulties (see *Figure 4*). In addition, imitation skills were strongly positively correlated with the composite score of cognitive skills at baseline, as well as communication skills (Receptive and Expressive Language) (ELC : *r*^*s*^ = 0.671, *p* < .001, RL : *r*^*s*^ *=* 0.778, *p* < .001, EL : *r*^*s*^ *=* 0.738, *p* < .001), that is, children with lower imitation skills were also those with lower skills in these domains (see *Figure 5*).

**Figure 4.**
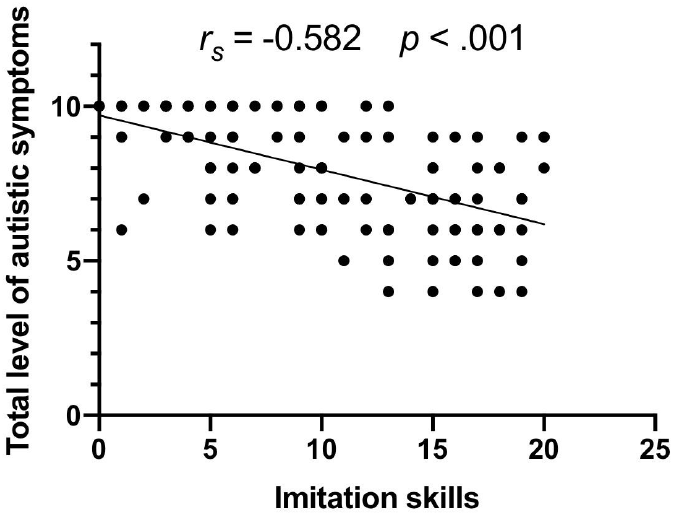
Spearman correlation between imitation and level of symptoms within the ASD group (Visuo-Motor Imitation scale of PEP-3 and Calibrated Severity Score of ADOS-G or 2)

**Figure 5.**
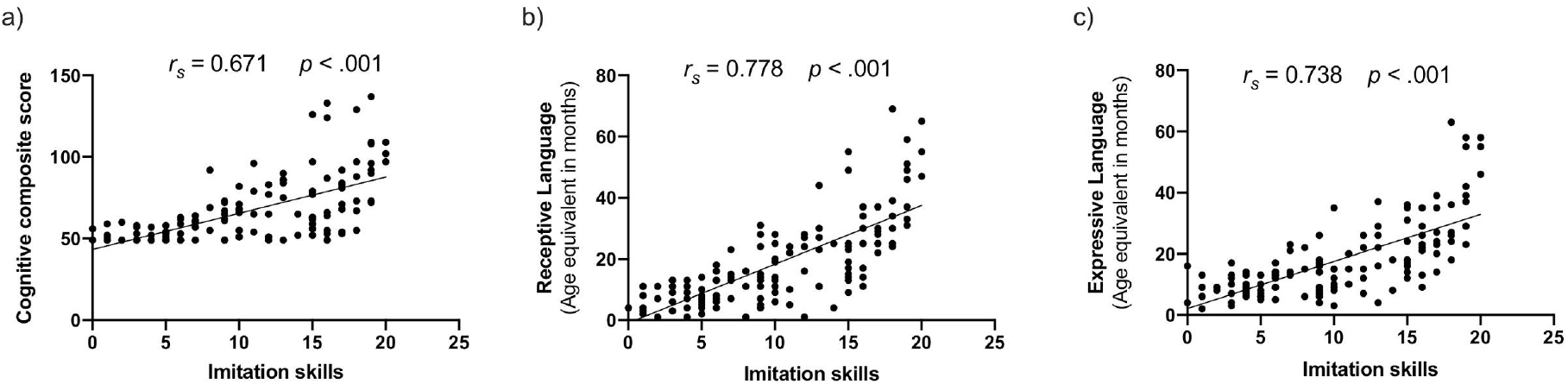
Spearman correlations between baseline imitation skills (raw scores of Visuo-Motor Imitation scale of PEP-3) and (a) Early Learning Composite of MSEL, (b) Receptive language age equivalent scores and (c) Expressive language age equivalent scores.

### Imitation at baseline as a predictor of the autistic symptoms and developmental changes

Imitation skills at baseline were predictive of the gains in communication skills (Receptive and Expressive Language) a year later in our preschoolers with ASD (RL : *r*^*s*^ = 0.371, *p* = .001, EL : *r*^*s*^ = 0.601, *p* < .001). Indeed, children with better imitation skills at baseline showed a greater improvement in these domains a year later (see *Figure 6*).This improvement was the most evident in the domain of expressive language. Further correlations between imitations skills and the composite cognitive score or the level of symptoms did not survive after correction for multiple comparisons (considering a stringent significance level at *p* < .0125 using Bonferroni correction).

**Figure 6.**
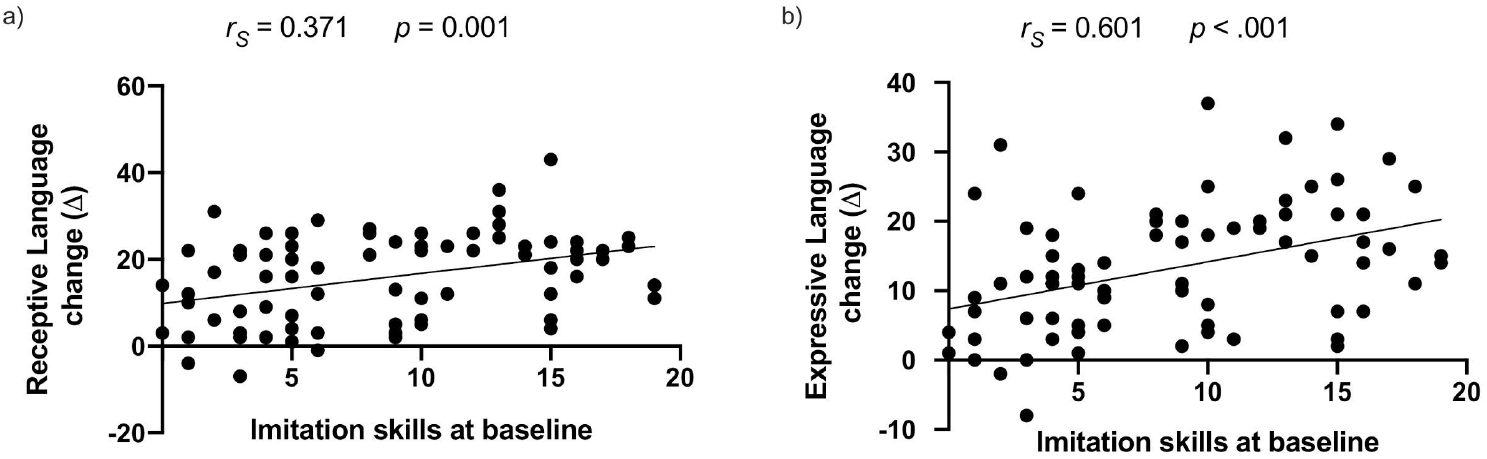
Spearman partial correlations between imitation skills (raw scores of the Visuo-Motor Imitation scale of PEP-3) and a one year gain in the ASD group for receptive language (a) and expressive language (b) (difference in age equivalent scores of the Mullen Scale of Early Learning).

### Imitation trajectories within the ASD group

The first step of K-means clustering on the baseline raw imitation scores yielded two optimal clusters (silhouette score of 0.828). The first cluster (sub-group) of ASD children (named ASD1, *n* = 46) (see *Figure 7)* had less difficulties with imitation at baseline and followed a trajectory highly similar to the one seen in our TD group (*Figure 3*) while the other cluster started significantly lower at baseline.

**Figure 7.**
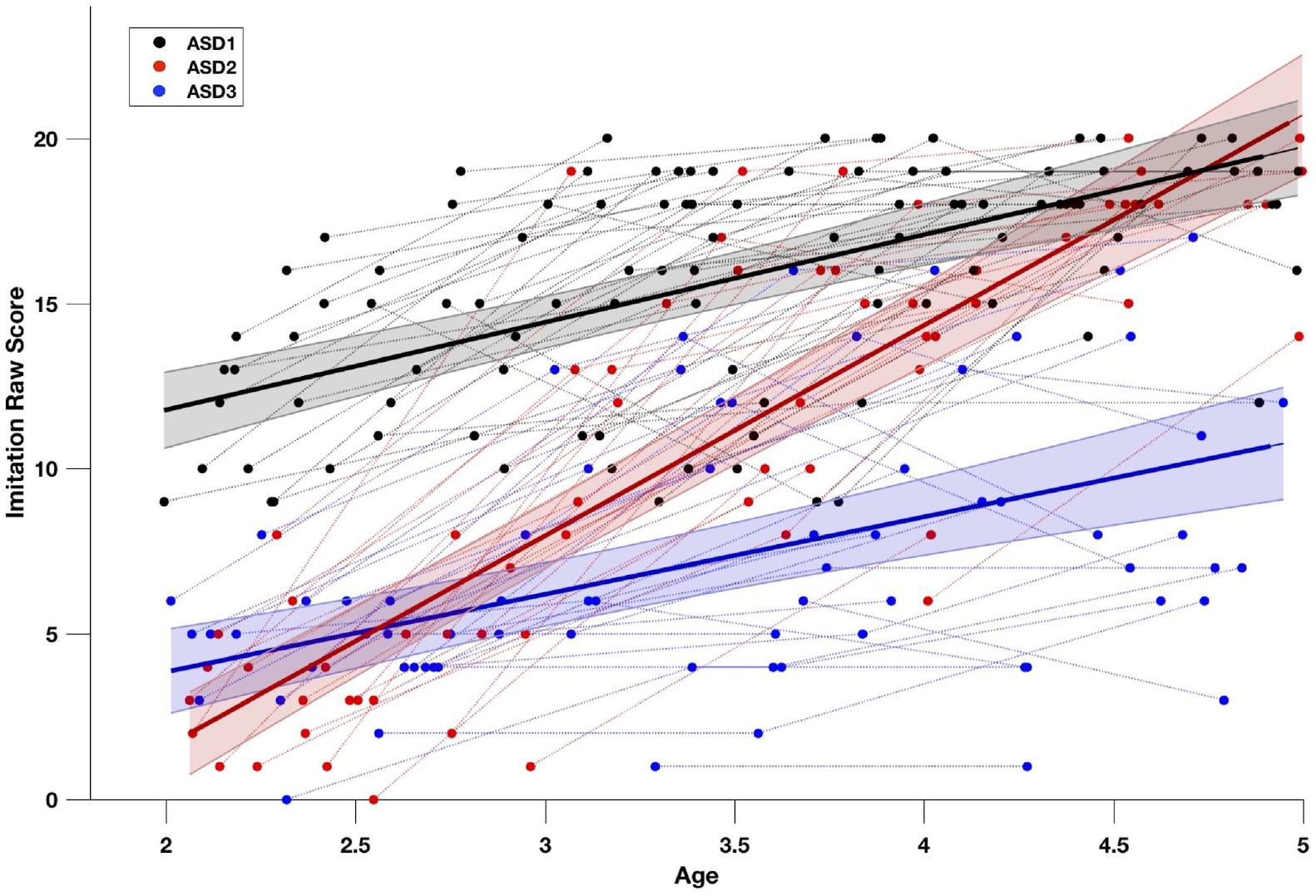
Imitation trajectories over time within the ASD group (*n* = 102). The trajectories at the group level (solid lines black=ASD1, *n*_*ASD1*_ = 46, red=ASD2, *n*_*ASD2*_ = 27, blue=ASD3, *n*_*ASD3*_ = 29) are obtained using mixed-effect models (Mancini & al., 2019; Mutlu & al., 2013). The 95% confidence interval of estimated group-level trajectory is represented in colored bands. Repeated time-points of the same subjects are connected with dotted lines. The distribution in the sub-groups was defined by the K-means clustering method in two steps. First clustering step yielded two clusters based on imitation skills raw scores at first time-point. We retained the higher cluster (ASD1), and proceeded to the second clustering step performed on the slopes of imitation skills, yielding ASD2 and ASD3 subgroups.

To better understand developmental trajectory in children who start lower on imitation skills, we then applied a second K-means clustering on the developmental slopes on imitation scores in our remaining group of ASD children. These children belonged to the cluster that showed less imitation skills at baseline (Total -ASD1 = 56 participants). This second clustering step yielded the two groups which we denote ASD2 and ASD3 (*n*^*ASD2*^ = 27, *n*^*ASD3*^ = 29). The trajectories of the two clusters from the second clustering step were significantly different (see *Figure 7*), one subgroup (ASD2) had a deficit on imitation skills at two years old, but showed a great improvement until five years old while the other subgroup in ASD children (ASD3) showed many difficulties in imitation at two years old and these difficulties continue over the time (group effect : *p* < .001 ; interaction : *p* < .001).

Finally, we wanted to understand which clinical characteristics were related to the distinct imitation trajectories in our sub-samples ASD2 and ASD3. In particular, we tested the baseline differences between the two groups (ASD2 and ASD3) with regards to the level of symptoms, cognitive and language skills, type of intervention and social orientation measured with an eye-tracking task (Franchini et al., 2017) (see *Table S2* in Supplementary Materials). We found no significant baseline differences between the ASD2 and the ASD3 according to all tested clinical variables. However, by analyzing these two groups’ trajectories in terms of many other aspects (communication skills, cognitive skills and adaptive behaviors), we observed that the subgroup ASD2 showed an overall improvement in all domains. This pattern suggests that imitation improvement in this group indicates an overall enhancement rather than the specific link of imitation skills progress and a particular developmental measure.

## Discussion

Our results corroborate previous findings on the imitative deficit in preschoolers with Autism Spectrum Disorder (ASD) in comparison with typically developing children (TD) (Colombi & al., 2009; Girardot & al., 2009; Ingersoll, 2008; McDuffie & al., 2007; Plavnick & Hume, 2014; Rogers & al., 1996; Thurm & al., 2007; Vivanti & Hamilton, 2014; Zwaigenbaum & al., 2005). Compared with our TD group, preschoolers with ASD showed significantly lower imitation skills as measured by the Visuo-Motor Imitation scale of the PEP-3 (Schopler & al., 2005). We then explored the trajectory of imitation skills over time, which is poorly documented in the literature until now (Young & al., 2011). Our results showed that TD children present good imitation skills, as measured with the PEP-3, already at the age of 2 years old, with only a slight improvement thereafter. In comparison, children with ASD presented a major imitative deficit at two years old, with a considerable gain up to five years old. We further confirmed that imitation skills in children with ASD at baseline were negatively related to the overall levels of symptoms while showing a positive relation with cognitive skills and language. We also showed that early imitation skills predicted the gains in communication skills one year after. Finally, we were able to delineate three different imitation trajectories in our sample of children with ASD. A subgroup of children (ASD1) presented fewer difficulties in imitation at two years old. In contrast, others (ASD2) showed major deficits in imitation at 2 years old, followed by accelerated gains up to the age of five. Lastly, we also identified a subgroup of children (ASD3) who had significant difficulties at baseline and showed less progress over the years.

### Developmental trajectory of imitation in ASD and TD

Besides showing lower baseline imitative skills, we found that the acquisition of imitation skills in children with ASD did not follow the same trajectory as in TD children. In ASD children, we observed the steep progress between the age of 2 and 3.5 years, highlighting the importance of this early sensitive period for learning new skills (Dawson, 2008; Guralnick, 1997; Johnson & Munakata, 2005; Lovaas, 1987; Vivanti et al., 2016). While most TD children reach the ceiling on this measure around the age of 5 years, the ASD group showed a lower average skill level at this age. Previous studies have suggested that the imitation deficits in ASD are more specific than broad (Rogers & Williams, 2006), concerning for example, more meaningless gestures than actions on the objects. Nevertheless, our results speak in favor of a more broad impairment as most of the actions measured by the imitation scale we used are rather simple one-step actions on objects and relatively few concern more abstract meaningless gestures. These difficulties in imitation of simple actions with objects can have deleterious effects on learning (Ingersoll, 2008; Ingersoll & Meyer, 2011; Poon et al., 2012; Villalobos et al., 2005). Indeed, our results showed that imitation skills were related to composite cognitive skills and communication skills (receptive and expressive language) in toddlers with ASD. As proposed by several authors (e.g. Wadsworth & al., 2017) a child with major imitative difficulties shows delay in the acquisition of new skills, especially because he/she does not use this channel to learn from her/his parents, siblings and peers (e.g. Howe & al., 2018; Zmyj & Seehagen, 2013).

In addition, confirming previous studies (McDuffie et al., 2007; Rogers et al., 2003; Zachor et al., 2010), our results showed a strong correlation between imitation skills and the level of symptoms measured by the ADOS-2 (Lord & al., 2012), whereby the children with more difficulties in imitation had also more symptoms of ASD at baseline. Thus, imitation skills were clinically informative of the severity of the level of autistic symptoms. This result corroborates findings claiming the importance of imitation in the acquisition of early tools of communication and its contribution to autistic symptoms (Hanika & Boyer, 2019; Ingersoll, 2008; Toth et al., 2006; Villalobos et al., 2005). Indeed, difficulties of engagement in reciprocal imitation early on may greatly limit social learning opportunities, lower the experiences of sharing and joint attention, important for socio-communicative development (Franchini et al., 2017). Moreover, taking part in imitation activities prepares the first basis of communication turns. Thus from the earliest moments, the reciprocal imitation of vocalisations and facial expressions in face-to-face interactions paves the way for the development of a more complex social interchange (Trevarthen et al., 1999, as cited by Ingersoll, 2008).

### Imitation as a predictor of developmental domains in children with ASD

While previous studies demonstrated that imitation predicts future cognitive skills (Hurley & Chater, 2005; Rogers & Pennington, 1991; Strid et al., 2006; Vivanti et al., 2013), the correlation that we observed in our sample did not hold after Bonferroni correction. The same was true for the relationship between imitation and change in the severity of autistics symptoms. Nevertheless, the baseline imitation skills were predictive of communication development one year later. Especially, gains in expressive language were greater in children with better imitation skills at baseline. The link between imitation and language has already been explored in previous studies (Heimann & al., 2016; Stone & al., 1997; Stone & Yoder, 2001; Toth & al., 2006). Indeed, infants start by imitating vocalizations, exchanges of vocalizations, then later words and sentences (e.g. Jones, 2007). Some studies found that verbal imitation (Smith et al., 2007) but also body movements imitation (Stone et al., 1997) are predictors of expressive language development.

Although children with ASD at a group level showed important gains in imitation skills, between two and five years old, the early difficulties in this domain presented at the age of two could still have an important influence on learning later on. Imitation is important not only for children’s current achievements, but also for their future skills. Learning to imitate can then be considered as one of the primary goals in early intervention programs for children with ASD (Rogers & Dawson, 2010), which can then be used to build on this skill for future acquisitions.

### Imitation trajectories within the ASD group

Given the important heterogeneity of the ASD phenotype (Wiggins et al., 2012), we explored the extent to which there might be different developmental trajectories of imitation within our sample of children with ASD. Although some authors have studied the trajectories of different types of imitation in children with ASD (Sanefuji & Yamamoto, 2014), to the best of our knowledge, no study to date tried to parse the heterogeneity of the early imitation development in ASD. Here, using a clustering approach, we observed three separate imitation trajectories in children with ASD. Similar to previous studies (Charman et al., 2003; Stone et al., 1997; Stone & Yoder, 2001; Vivanti et al., 2013), we found that good early imitation skills are linked to better outcomes, while early difficulties predict impairment in developmental domains. Nevertheless, our study showed that some children with ASD have major difficulties in imitation at 2 years old, but show a great improvement until 5 years of age. As mentioned previously, we were not able to find a characteristic that would differentiate these two groups at baseline and predict whether a child with imitation difficulties at baseline will follow one trajectory or the other. However, our results support that imitation improvement is an indicator of an overall enhancement, without a clear causal relationship with the used measures. More investigation should be done to understand the mechanisms through which a child’s development is set on one trajectory or the other. The absence of evidence for specific differences between the two groups with a divergent imitation trajectory is interpreted with caution. Indeed, imitation is often used to incite and encourage the child to do the exercises in assessments such as the one used in this study (MSEL; Mullen, 1995), specifically when there are difficulties of language comprehension, as it is frequently the case in young children with ASD (Kwok et al., 2015). Thus, a low score on domains of cognitive or language skills could, in some cases, be better explained by maladaptive behaviors than by real difficulties in these areas (Robain et al., 2020).

### Perspectives and limitations

As a limitation to this study, we would like to emphasize that the imitation skills measured in the context of our study are elementary skills, and concern mainly actions with objects. Thus, it would be particularly interesting to refine the analyses using a scale adapted for a larger age range, with a broader variety of activities, allowing differentiation of different imitation types as described by Vivanti & Hamilton (2014).

There are many avenues to pursue to better understand the mechanisms underlying imitation in young children with ASD. Indeed, as discussed, it would be particularly interesting and valuable to find an early predictor of the developmental trajectory in children with early imitation difficulties. We showed that the children with an imitation deficit at 2 years will either maintain their difficulties in a rather global way in the years to come, or show significant improvement in imitation but also in other areas in general. It would then be a question of finding which characteristics of the behavioral phenotype, or the interventions received allowed the positive evolution in some cases. One of the avenues to follow in this sense could be to explore earlier milestones of imitation than one measured in our study. Indeed, imitation is very linked to other skills such as social orientation, joint attention and social reciprocity as demonstrated by some authors (Girardot et al., 2009; McDuffie et al., 2007). Despite our unsuccessful attempt to distinguish our two groups in terms of social orientation using an eye-tacking task (Franchini et al., 2017), more investigations could be done in this sense with a younger sample.

## Conclusions

Our study confirmed a delay in the acquisition of imitation skills in preschoolers with ASD who present a different developmental trajectory than children with typical development. We also showed that these difficulties are related to levels of autistic symptoms, developmental deficits, and long-term consequences on the development of communication and composite cognitive skills. Moreover, our analyses identified subgroups of children with ASD in terms of imitative development, reflecting the heterogeneity that characterizes this population. Thus, our study confirms the status of imitation as a prerequisite for the development of other skill areas and the importance of including its learning in early intervention programs, for example.

## Data Availability

Following the publication in a peer reviewed journal, the data used in this study can be obtained upon a reasonable request.

## Acknowledgements

The authors are particularly grateful to all the families who participated in this study. Furthermore, the authors thank Lylia Ben Hadid, François Robain, Sonia Richetin, Sara Maglio, Aurélie Bochet, Michel Godel, Stefania Solazzo, Ornella Vico Begara, Chiara Usuelli, Eva Micol, Laura Sallin, Lisa Esposito, Kenza Latrèche, Oriane Grosvernier and Constance Ferrat for their implication in data collection.

