## Supplementary Materials for "Trajectories of imitation skills in preschoolers with Autism Spectrum Disorders"

### Statistical comparison between groups at baseline

As our baseline distributions didn't follow normal distributions, the statistical comparisons between groups at baseline were on medians which were presented in *Table S1*.

**Table S1.** *Statistical Comparison Between Children With ASD and Children with TD at Baseline in Terms of Demographic, Clinical, Cognitive Features*

| | ASD - *Mdn n* = 177 | TD - *Mdn n* = 43 | *p*-value* |
|---|---|---|---|
| Sex | 24 ♀ / 153 ♂ | 10 ♀ / 33 ♂ | 0.155[1] |
| Age (years old) | 2.82 | 2.76 | 0.429[2] |
| ADOS Total symptom severity | **8** | **1** | **<0.001**[2] |
| PEP-3 | | | |
| CVP (raw score) | **27** | **45** | **<0.001**[2] |
| EL (raw score) | **4** | **27** | **<0.001**[2] |
| RL (raw score) | **10** | **32** | **<0.001**[2] |
| VABS-II Adaptive Behavior Composite | **77** | **110** | **<0.001**[2] |

*Note.* ADOS = Autism Diagnosis Observation Schedule ; PEP-3 = PsychoEducational Profile, 3rd edition ; CVP = Cognition Verbal and Preverbal ; EL = Expressive language ; RL = Receptive language ; VABS-II = Vineland Adaptive Behavior Scales, 2nd edition.
* p value of *Fisher's exact test*[1] and *Mann-Whitney tests*[2] on medians of differences between the ASD and TD groups. Significant results are shown in bold.

### Statistical comparison between ASD subgroups at baseline

In order to differentiate the subgroups ASD2 and ASD3 at baseline, we compared them according to the level of symptoms (ADOS Total symptom severity) and cognitive and language skills (PEP-3). In addition, we compared the number of children in each group that was included in an early and intensive Early Start Denver Model intervention program in Geneva (Rogers & Dawson, 2010). Finally, in order to test for potential differences between the two groups with regards to social orienting, we used eye-tacking. In this task adynamic social and geometric videos are presented side-by-side (see Franchini et al., 2017, for more details). Thus, as a measure of social orienting, for each child we were able to calculate the percentage of time she/he spent on the videos containing social information compared to the videos with geometric forms .

**Table S2.** *Statistical Comparison Between Subgroups ASD2 and ASD3 at Baseline in Terms of Clinical, Cognitive, Therapy and Eye-tracking Features*

| | ADS2 - *Mean (SD)* $n = 27$ | ASD3 - *Mean (SD)* $n = 29$ | $p$-value |
|---|---|---|---|
| ADOS Total symptom severity | 8.46 (1.6) | 9.07 (1.2) | $p = 0.179$[1] |
| PEP-3 | | | |
| VPC (raw scores) | 15.81 (8) | 13.61 (6.7) | $p = 0.613$[1] |
| EL (raw scores) | 2.96 (4.7) | 1.71 (1.2) | $p = 0.613$[1] |
| RL (raw scores) | 4.73 (6) | 3.71 (3) | $p = 0.873$[1] |
| Number of subjects included in an early intervention (CIPA) | 15 | 13 | $p = 0.408$[2] |
| Social orientation (eye-tracking) | 39.89 (19.3) | 45.68 (19) | $p = 0.937$[3] |

*Note.* ADOS = Autism Diagnosis Observation Schedule ; PEP-3 = PsychoEducational Profile, 3rd edition ; CVP = Cognition Verbal and Preverbal ; EL = Expressive language ; RL = Receptive language.
* p value of *Mann-Whitney tests*[1] , *Chi-square test*[2] and *t-test*[3] of differences between the two subgroups.
